# Standard precautions perception and practice among health workers in the obstetrics-gynecology department of a referral hospital in Cameroon

**DOI:** 10.1101/2025.04.21.25326141

**Authors:** Fabrice Zobel Lekeumo Cheuyem, Emilia Enjema Lyonga, Innocent Takougang

## Abstract

**Background:** Standard precautions are infection control and prevention (ICP) measures designed to reduce the risk of transmission of infectious agents in healthcare settings. In Cameroon, maternal and neonatal mortality remain a concern. Unsystematic compliance with these measures increases the risk of HAIs and other medical hazards. The present investigation aimed at assessing the baseline understanding of hand hygiene principles and perceptions, experience of occupational exposure to body fluids, and preventive vaccination coverage among HCWs in the obstetrics-gynecology ward of a referral hospital of Yaounde.

**Methods:** A descriptive cross-sectional study was conducted in the obstetrics-gynecology department of the health facility from April to July 2024. A self-administered questionnaire was used to collect data from consenting HCWs. It was adapted from the WHO Knowledge Questionnaire for Health Care Workers.

**Results:** A total of 41 healthcare workers participated in the study. Participants were predominantly female (78%) and ranged in age from 20 to 57 years. Staff knowledge of hand hygiene was average overall, with a median score of 60%. Most of respondents considered hand hygiene to be an essential part of their care (90%). Most of paramedical staff (82%) significantly agreed that they had been properly trained on hand hygiene during their training (*p*=0.006). More than half of respondents (59%) experienced an accidental exposure to body fluids in the previous 12 months. A significant factor associated with the occurrence of occupational exposure to blood and other body fluids was a high level of education (aOR=14; *p*=0.044). The coverage of fully vaccinated HCWs was 27% for viral hepatitis B, 19% for COVID-19, and 0% for cholera. Factors associated with low adherence to vaccination included not having received training in ICP interventions (aOR=7.37; *p*=0.046) for hepatitis B vaccination and having completed tertiary education (aOR=14; *p*=0.043) for COVID-19. Half of the HCWs exposed (12/24=50%) to blood and body fluids were not fully vaccinated against viral hepatitis B.

**Conclusions:** This study revealed gaps in knowledge of hand hygiene, high occupational exposure to biological fluids and low vaccination. Health facility managers and national health authorities must therefore commit to implementing specific strategies to increase staff training in standard precautions and promote vaccination.

## Background

Standard precautions are infection control and prevention (ICP) measures designed to reduce the risk of transmission of infectious agents from recognized and unrecognized sources to healthcare workers (HCWs), patients, and visitors in healthcare settings [1]. Hand hygiene is a critical component of standard precautions and offers a simple yet effective solution to reducing healthcare-associated infections (HCAIs) and the spread of antimicrobial resistance across various healthcare settings, from advanced systems to primary healthcare centers [2,3]. Unsystematic compliance with these measures increases the risk of HAIs and other medical hazards [1].

The global healthcare landscape is compromised by the high incidence of HAIs, which are among the leading adverse events in healthcare settings [4]. Globally, nearly 9 million HAIs occur each year, resulting in approximately 1 million deaths with low- and middle-income countries bearing a disproportionate impact of this burden [5,7,8].

In Cameroon, the burden of occupational exposure to body fluids is high regardless of the health facility category. The prevalence among HCWs varies from 20 to 80%, largely due to poor adherence to standard precautions [8–10]. This exposure can lead to serious and potentially life-threatening diseases including viral hepatis B, COVID-19, cholera, measles, varicella and other pathogens transmitted through blood and other body fluids [11].

One of the most cost-effective strategies to prevent these infections is the vaccination of HCWs [12,13]. However, health authorities face several barriers to making to this vaccine fully accessible to HCWs [14]. Such barriers, except the economic accessibility, include vaccine hesitancy which leads to the refusal or delayed acceptance of vaccines despite the availability of immunization services [15,16]. Study reports highlight various factors that contribute to this hesitancy, including perceptions of disease risk and severity, mistrust of vaccines, fear of needles and adverse events, vaccines cost, societal factors such as social norms and religion [9,14,15]. These concerns have been fueled by the emergence of the COVID-19 and the rapid development of preventive vaccines [14,16].

High maternal (596/100,000 live births) and neonatal (25.6/1,000 live births) mortality persist in the country [17,18]. The primary drivers of maternal mortality include sepsis, abortion, place of delivery, and suboptimal quality of care [19]. Inadequate delivery services result in significant puerperal (9-17%) and post-caesarean wound (2-32%) infections across health facilities [20–22].

Adherence to infection prevention measures by HCWs during labor and delivery has long been recognized as an important strategy for mitigating infection risks [23]. However, compliance with hand hygiene and other infection prevention measures among HCWs remains suboptimal among. To address this issue, sustained efforts are needed to identify and implement effective strategies. Education of HCWs and healthcare trainees is a key strategy in this endeavor [2,24].

To support data-driven decision-making, an assessment of health facilities providing maternal and neonatal care was needed. The objective of this investigation was to assess the baseline understanding of hand hygiene principles and perceptions, prevalence of occupational exposure to body fluids, and preventive vaccination coverage among HCWs in the obstetrics-gynecology ward of a referral hospital of Yaounde.

## Methods

A descriptive cross-sectional research design was used to collect data over a three-month period (May-July 2024) in a referral hospital of Yaounde.

Yaounde is It is a city of approximately 3.2 million inhabitants covered by first, second, third and fourth level health facilities. Yaounde is the political capital of Cameroon with a population of around 3.2 million. This study setting is a leading public health institution. It serves as the fourth level of reference in the national health pyramid, providing specialized health services. It has a capacity of 138 beds and receives a significant number of patients, with 500 to 1,000 admissions per week [25–27].

A convenience sampling approach was used to enroll all consenting participants among the 48 HCWs present in the obstetrics-gynecology ward during the study period. They were residents, nurses, midwives/obstetric nurses, nursing assistants, and students.

A pre-tested questionnaire addressed to HCWs It was adapted from the knowledge assessment tool developed by WHO [2,28]. It included questions with items related to demographic characteristics, hand hygiene, occupational exposure to body fluids, vaccination against hepatitis B, COVID-19 and cholera.

A score of 1 was assigned for each correct response to the hand hygiene knowledge assessment and the score classified into three categories: good (≥75%), average (50-74%) and poor (<50%) [29]. Data were checked, coded and analyzed using R Statistics version 4.3.3. Fisher’s probability test was used to compare proportions. The QQ diagram was used to compare the distribution of quantitative variables with respect to the normal distribution. The nonparametric Kruskal-Wallis’s test was used to compare median scores between three or more study groups.

The student variable represented those undergoing training in general medicine who were on placement in the gynecology-obstetrics department. The term resident referred to students undergoing specialization in gynecology-obstetrics unit. The professional group variable included medical staff represented by residents, paramedical professionals (nurses, midwives/obstetric nurses, assistant-nurses) and medical students.

Participants were considered fully vaccinated if they had received at least three doses of hepatitis B vaccine according to the recommended schedule. For the COVID-19 vaccine, anyone who had received at least one dose of the Janssen vaccine or at least two doses of the Sinopharm or AstraZeneca vaccine was considered fully vaccinated.

Simple and multiple binary logistic regressions were used to assess the strength of association between variables and to remove potential confounders. The best predictors for the model were progressively selected using Akaike’s information criterion (AIC). Confidence intervals (CIs) were estimated at the 95% level. A *p*-value<0.05 was considered statistically significant.

This study received ethical clearance from the Institutional Research Ethics Committee (CIER) of the Faculty of Medicine and Biomedical Sciences, University of Yaounde 1, under protocol number 1017/UYI/FMSB/VDRC/DAASR/CSD. Additional approval was obtained from the General Management of the health facility. Written informed consent was obtained from all participants prior to their enrolment in the study. All procedures adhered strictly to the tenets of the Declaration of Helsinki.

## Results

Among 47 contacted HCWs, 46 (98%) participated, with 41 (89%) returning fully completed questionnaires.

### Socio-professional characteristics of study participants

Participants ranged in age from 20 to 57 years, with a median age of 30 [22; 47] years. They were predominantly female (78%) and had higher education (73.2%). Their median work experience was 3 [3; 19] years.

### Hand hygiene knowledge

Nearly two thirds of respondents (68%) identified hand hygiene before touching a patient as the ideal time to prevent the transmission of germs. However, less than a third (20%) had poor knowledge of the ideal time for this practice. Less than a third of HCWs (20%) had good knowledge of the minimum duration of action of hydroalcoholic gel. Medical staff (63%) had significantly better knowledge (*p*=0.027) (Table 1).

**Table 1.** Hand hygiene knowledge of healthcare workers in the gynecology-obstetrics department, May 2024 (*n*=41)

| Assessed knowledge | Total | Professional group |  |  | *p*-value<sup>1</sup> |
| --- | --- | --- | --- | --- | --- |
|  |  | *n* (%) |  |  |  |
|  |  | Medical<br>8 (100) | Paramedical<br>23 (100) | Student<br>10 (100) |  |
| **Timing of hand hygiene to prevent transmission of germs to the patient** |  |  |  |  |  |
| Immediately before a clean/aseptic procedure (*Yes*) | 12 (29) | 2 (25) | 7 (30) | 3 (30) | >0.999 |
| Immediately after exposure to body fluids (*Yes*) | 12 (29) | 2 (25) | 7 (30.4) | 3 (30) | >0.999 |
| Before touching a patient (*Yes*) | 28 (68) | 7 (88) | 13 (57) | 8 (80) | 0.202 |
| After exposure to the patient's immediate environment (*No*) | 8 (20) | 3 (38) | 3 (13) | 2 (2) | 0.322 |
| **Minimum duration of efficacy of hydroalcoholic gel (20 sec)** | 10 (24) | 5 (63) | 4 (17) | 1 (10) | **0.027** |
| **Elements associated with an increased probability of germ colonization of hands** |  |  |  |  |  |
| Injured skin (Yes) | 24 (59) | 4 (50) | 13 (57) | 7 (70) | 0.755 |
| Wearing jewelry (Yes) | 20 (49) | 3 (38) | 10 (43) | 1 (10) | 0.214 |
| Artificial nails (Yes) | 26 (63) | 5 (63) | 16 (70) | 5 (50) | 0.564 |
| Regular use of hand cream (No) | 2 (5) | 0 (0) | 2 (9) | 0 (0) | >0.999 |
| **Best method of hygiene in each situation** |  |  |  |  |  |
| Before palpating an abdomen (Hydroalcoholic gel) | 26 (63) | 6 (75) | 14 (61) | 6 (60) | 0.822 |
| After removing examination gloves (Washing) | 31 (76) | 7 (86) | 17 (74) | 7 (70) | 0.785 |
| Before administering an injection (Hydro-alcoholic gel) | 19 (46) | 5 (63) | 9 (39) | 5 (50) | 0.534 |
| After emptying a bedpan (Washing) | 36 (88) | 7 (88) | 20 (87) | 9 (90) | >0.999 |
| After making the patient's bed (Hydro-alcoholic gel) | 15 (37) | 1 (13) | 10 (43) | 4 (40) | 0.369 |
| After exposure to blood (Washing) | 37 (90) | 7 (88) | 21 (91) | 9 (90) | >0.999 |
| **Knowledge score** |  |  |  |  |  |
| Median score | 60 | 57 | 60 | 60 | 0.9<sup>(2)</sup> |
| Interquartile range | 53 - 67 | 53 - 63 | 50 - 67 | 48 - 65 |  |

<sup>1</sup> Fisher exact probability test; <sup>2</sup> Kruskal-Wallis's test

### Perception and practice of hand hygiene

Most of respondents (88%) reported that they always practiced hand hygiene and considered it an essential part of their care (90%). Most respondents (39%) disagreed with the statement that no hand hygiene courses were offered in their departments (Figure 1).

**Fig. 1.**
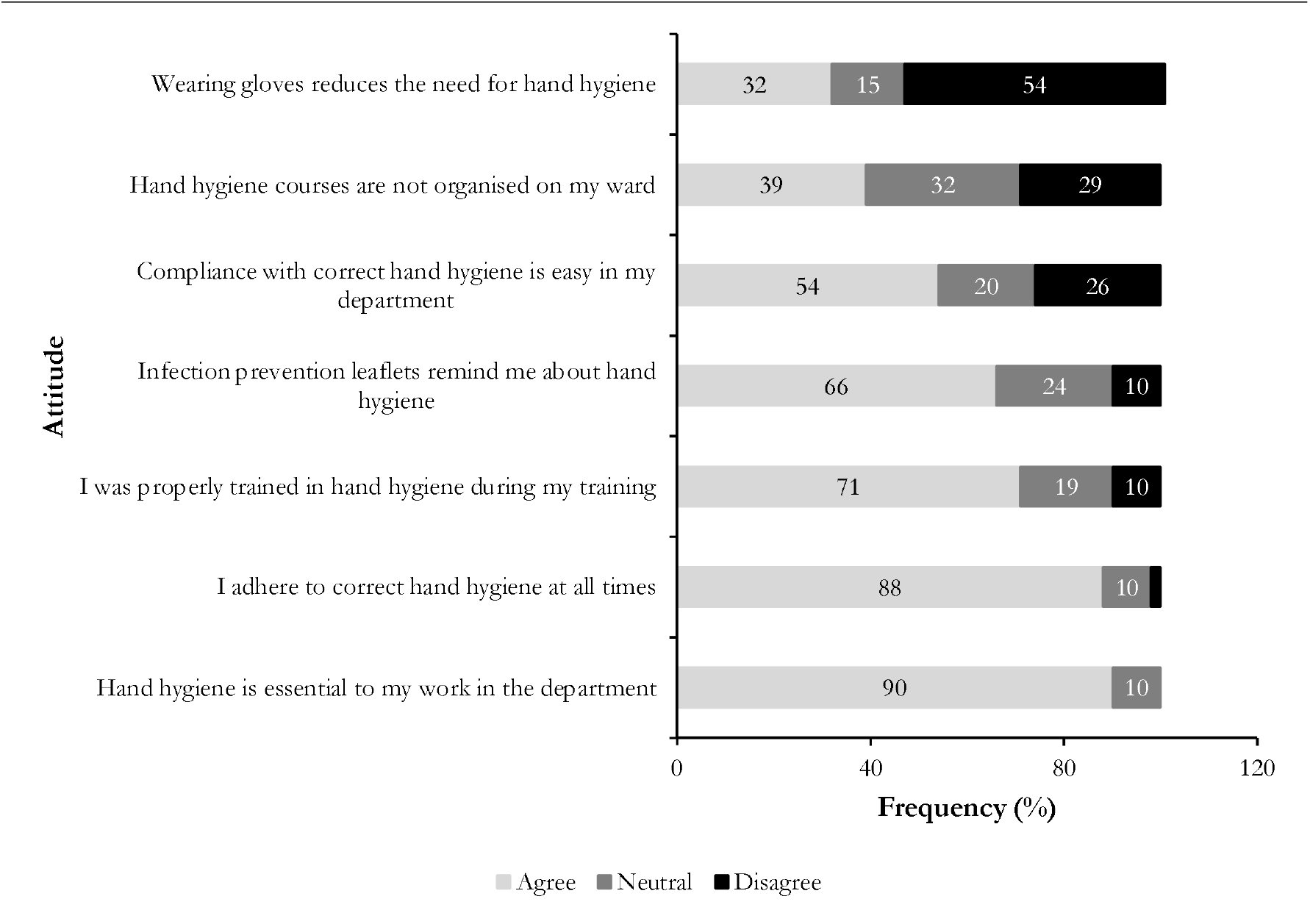
Attitude and practice related to hand hygiene among healthcare workers at the gynecology-obstetrics unit, May 2024 (*n* =41)

Most of paramedical staff (82%) significantly agreed that they had been properly trained on hand hygiene during their training (p=0.006). In contrast, medical students were less likely to agree with this statement (20%) (Table 2).

**Table 2.** Attitude and practice relating to hand hygiene according to professional group in the gynecology-obstetrics department, May 2024 (*n*=41)

| Attitude & Practice | Professional group<br>*n* (%) |  |  | *p*-value<sup>/</sup> |
| --- | --- | --- | --- | --- |
|  | Medical<br>8 (100) | Paramedical<br>23 (100) | Medical<br>8 (100) |  |
| **I adhere to correct hand hygiene at all times** |  |  |  | >0.999 |
| Agree | 7 (88) | 20 (87) | 9 (9) |  |
| Neutral | 1 (13) | 2 (9) | 1 (10) |  |
| Disagree | 0 (0) | 1 (4) | 0 (0) |  |
| **Wearing gloves reduces the need for hand hygiene** |  |  |  | 0.969 |
| Agree | 2 (25) | 8 (35) | 3 (30) |  |
| Neutral | 1 (13) | 3 (13) | 2 (20) |  |
| Disagree | 5 (62) | 12 (52) | 5 (50) |  |
| **I was properly trained in hand hygiene during my training** |  |  |  | 0.006 |
| Agree | 4 (50) | 21 (82) | 4 (40) |  |
| Neutral | 3 (37) | 1 (4) | 4 (40) |  |
| Disagree | 1 (13) | 1 (4) | 2 (20) |  |
| **Compliance with correct hand hygiene is easy in my department** |  |  |  | 0.154 |
| Agree | 2 (25) | 16 (70) | 4 (40) |  |
| Neutral | 2 (25) | 3 (13) | 3 (30) |  |
| Disagree | 4 (50) | 4 (17) | 3 (30) |  |
| **Hand hygiene is essential to my work in the department** |  |  |  | 0.625 |
| Agree | 7 (88) | 20 (87) | 10 (100) |  |
| Neutral | 1 (13) | 3 (13) | 0 (0) |  |
| **Infection control leaflets remind me about hand hygiene** |  |  |  | 0.851 |
| Agree | 5 (62) | 14 (61) | 8 (80) |  |
| Neutral | 2 (25) | 6 (26) | 2 (20) |  |
| Disagree | 1 (13) | 3 (13) | 0 (0) |  |
| **Hand hygiene courses are not organized in my unit** |  |  |  | 0.143 |
| Agree | 3 (37) | 9 (39) | 4 (40) |  |
| Neutral | 4 (50) | 4 (17) | 5 (50) |  |
| Disagree | 1 (13) | 10 (44) | 1 (10) |  |

<sup>/</sup> Fisher exact probability test

### Occupational exposure to body fluids

A total of 24 HCWs (59% of respondents) experienced an accidental exposure to biological fluids in the last 12 months. Splashes were the main cause of exposure (96% of cases), followed by percutaneous exposure (16%).

The biological fluids most frequently involved were blood (50%) and amniotic fluid (44%) (Figure 2).

**Fig. 2.**
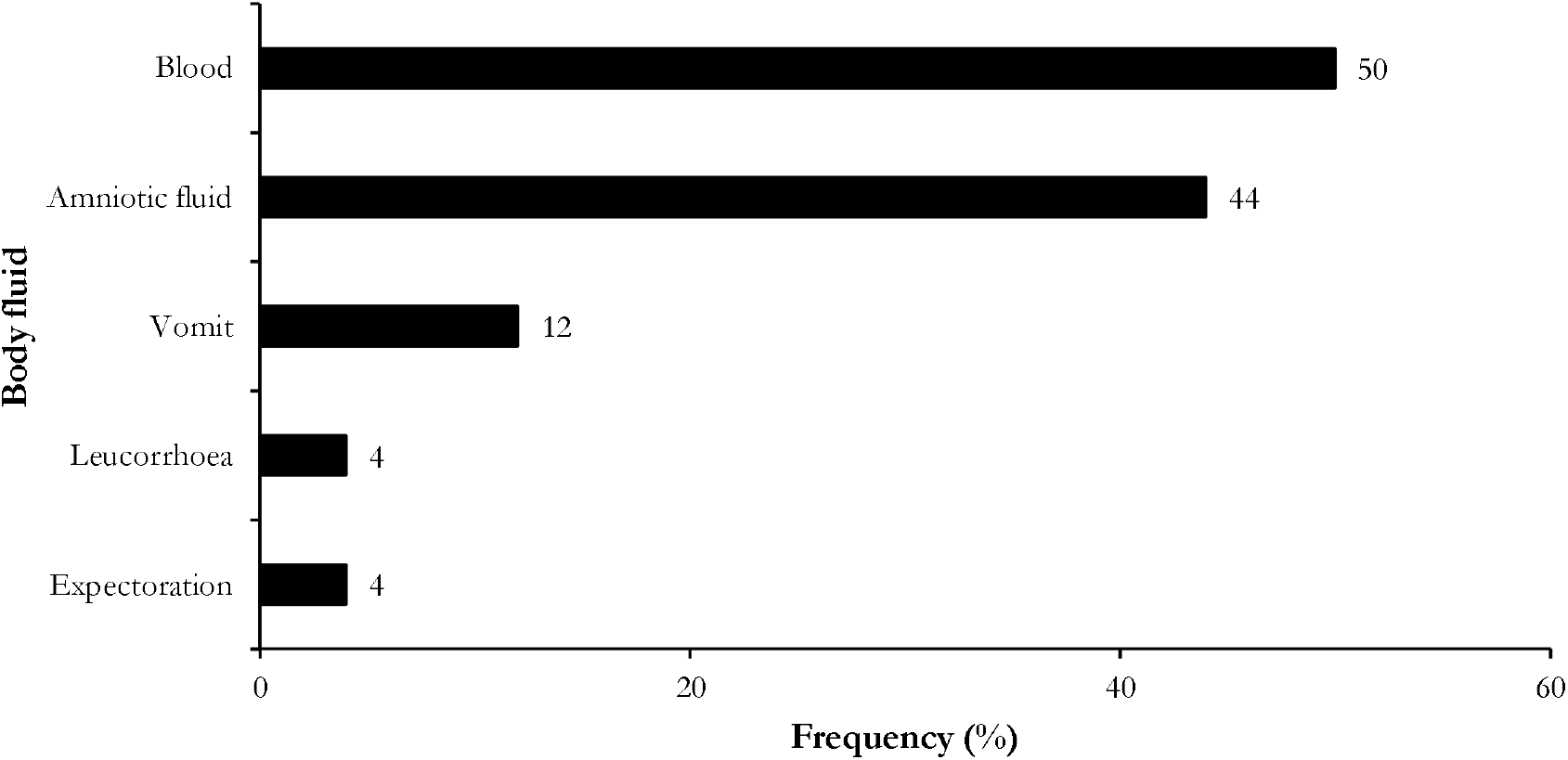
Reported types of biological fluids involved in occupational exposures by health care workers in the gynecology-obstetrics, May 2024 (*n*=24)

Univariate analysis revealed two factors significantly associated with occupational exposure to biological fluids: grade (OR=11; *p*=0.046) and hepatitis B vaccination status (OR=7.5; *p*=0.018). Multivariate analysis revealed that healthcare workers with a higher level of education were significantly 14 times more likely to be accidentally exposed to biological fluids than those with a secondary level of education (*p*=0.044).

In addition, half of the staff exposed (12/24=50%) to blood and body fluids were not fully vaccinated against viral hepatitis B (Table 3).

**Table 3.** Simple and multiple binomial logistic regressions of parameters associated with occupational exposure to biological fluids among healthcare workers in the gynecology-obstetrics, May 2024 (*n*=41)

| Factor | Exposition<br>*n* (%) |  | Total<br>*n* | cOR | *p*-value | aOR | 95% CI limits |  | *p*-value<sup>2</sup> |
| --- | --- | --- | --- | --- | --- | --- | --- | --- | --- |
|  | Yes<br>24 (58) | No<br>17 (42) |  |  |  |  | Lower | Upper |  |
| **Age<sup>1</sup>** |  |  |  |  |  |  |  |  |  |
| □25 | 8 (53) | 7 (47) | 15 | 1 |  |  |  |  |  |
| 25 - 34 | 8 (89) | 1 (11) | 9 | 7.0 | 0.099 |  |  |  |  |
| 35 - 44 | 2 (40) | 3 (60) | 5 | 0.6 | 0.607 |  |  |  |  |
| 45 + | 6 (50) | 6 (50) | 12 | 0.9 | 0.863 |  |  |  |  |
| **Gender** |  |  |  |  |  |  |  |  |  |
| Male | 5 (56) | 4 (44) | 9 | 1 |  | 1 |  |  |  |
| Female | 19 (59) | 13 (41) | 32 | 1.2 | 0.837 | 2.3 | 0.3 | 24.1 | 0.451 |
| **Marital status** |  |  |  |  |  |  |  |  |  |
| Single | 16 (55) | 13 (45) | 29 | 1 |  | 1 |  |  |  |
| Married | 8 (67) | 4 (33) | 12 | 1.6 | 0.498 | 1.9 | 0.3 | 16.3 | 0.520 |
| **Educational level** |  |  |  |  |  |  |  |  |  |
| Secondary | 4 (36) | 7 (64) | 11 | 1 |  | 1 |  |  |  |
| Higher | 20 (67) | 10 (33) | 30 | 3.5 | 0.089 | 14.2 | 1.1 | 503 | **0.044** |
| **Grade** |  |  |  |  |  |  |  |  |  |
| Nurse | 5 (38) | 8 (62) | 13 | 1 |  | 1 |  |  |  |
| Resident | 7 (87) | 1 (13) | 8 | 11.2 | **0.046** | 12.1 | 0.8 | 483 | 0.112 |
| Midwife/Obstetric nurse | 5 (71) | 2 (29) | 7 | 4.0 | 0.170 | 1.1 | 0.1 | 24.8 | 0.966 |
| Nurse-assistant | 2 (67) | 1 (33) | 3 | 3.2 | 0.389 | 9.4 | 0.3 | 596 | 0.214 |
| Student | 5 (50) | 5 (50) | 10 | 1.6 | 0.581 | 2.4 | 0.2 | 43.7 | 0.510 |
| **Professional group** |  |  |  |  |  |  |  |  |  |
| Student | 5 (50) | 5 (50) | 10 | 1 |  |  |  |  |  |
| Paramedical | 12 (52) | 11 (48) | 23 | 1.1 | 0.908 |  |  |  |  |
| Medical | 7 (87) | 1 (13) | 8 | 7.0 | 0.117 |  |  |  |  |
| **Professional experience 1<sup>1</sup>** |  |  |  |  |  |  |  |  |  |
| □5 | 15 (68) | 7 (32) | 22 | 1 |  |  |  |  |  |
| 5 + | 9 (47) | 10 (53) | 19 | 0.4 | 0.181 |  |  |  |  |
| **Professional experience 2<sup>1</sup>** |  |  |  |  |  |  |  |  |  |
| □10 | 16 (67) | 8 (33) | 24 | 1 |  |  |  |  |  |
| 10 + | 8 (47) | 9 (53) | 17 | 0.4 | 0.212 |  |  |  |  |
| **Hepatitis B vaccination status** |  |  |  |  |  |  |  |  |  |
| Unvaccinated | 12 (44) | 15 (56) | 27 | 1 |  |  |  |  |  |
| Vaccinated | 12 (86) | 2 (14) | 14 | 7.5 | **0.018** |  |  |  |  |

<sup>1</sup>Unit in years; <sup>2</sup>Fisher exact probability test; cOR: crude Odds Ratio; aOR: adjusted Odds Ratio; CI: Confidence interval

Most of HCWs (73%) reported that there were no posters on the ward about management of accidental exposures to blood and other biological fluids.

### Compliance with preventive vaccination

The coverage of fully vaccinated HCWs was 27% for hepatitis B and 19% for COVID-19. However, no participant was fully vaccinated against cholera, with a small proportion partially vaccinated (5%) (Figure 3).

**Fig. 3.**
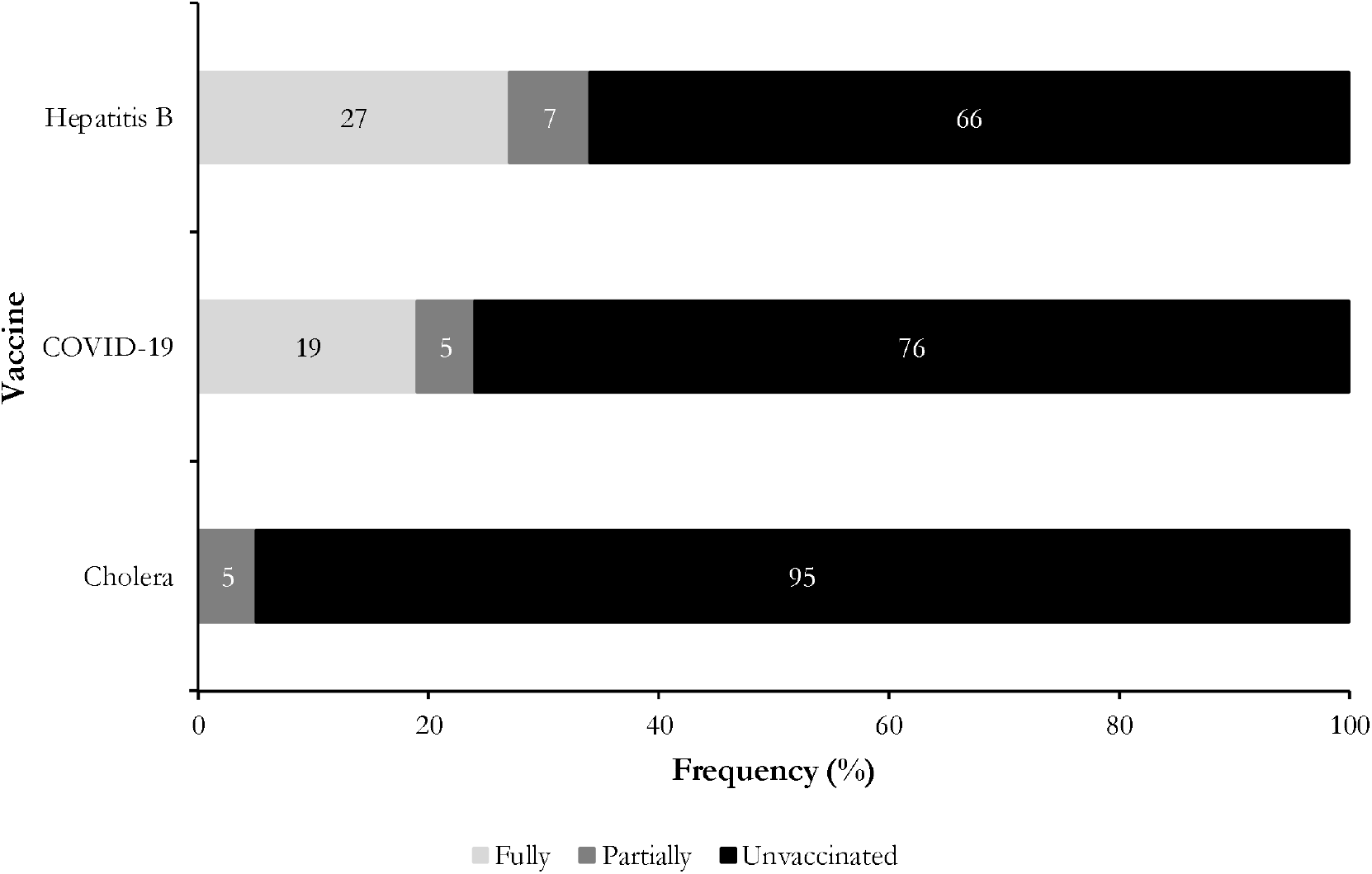
Vaccination coverage against viral hepatitis B, COVID-19 and cholera among health workers in the gynecology-obstetrics department, May 2024 (*n*=41)

### Hepatitis B vaccination

The results of the multivariate analysis showed that the following factors were associated with low adherence to viral hepatitis B vaccination:

Age: workers aged 45 and over were three times more likely not to be vaccinated than those aged under 25 (aOR=2.6).

Gender: Men were three times more likely not to be vaccinated than women (aOR=2.8).

Marital status: married workers were 40% more likely not to be vaccinated than unmarried workers.

Level of education: staff with a higher level of education were 50% more likely not to be vaccinated than those with a secondary level of education.

Training: staff who had not received training in ICP measures were significantly seven times more likely not to be vaccinated than those who reported having received such training (aOR=7.37; *p*=0.046) (Table 4).

**Table 4.**
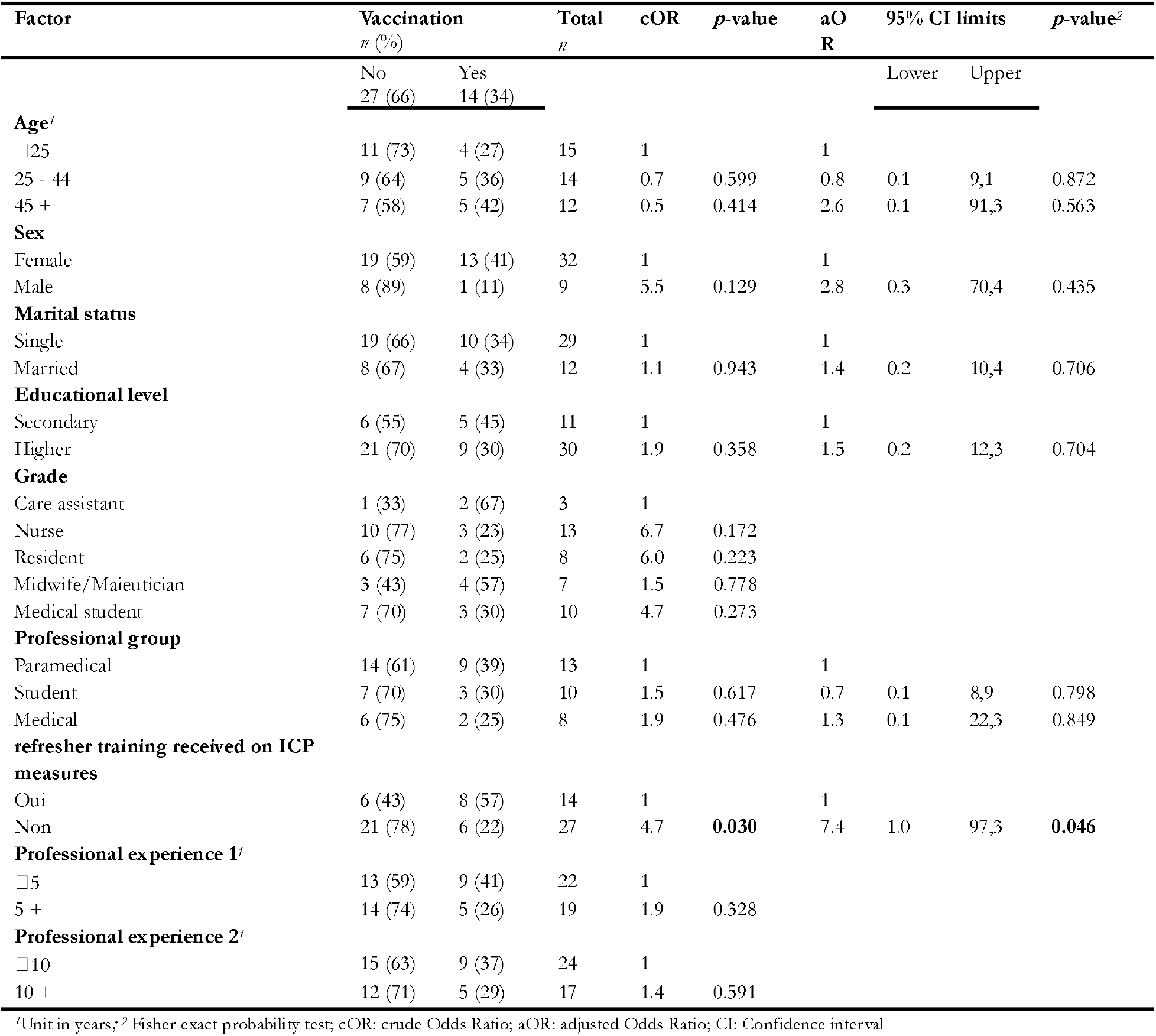
Univariate and multivariate analysis of parameters associated with non-adherence to vaccination against viral hepatitis B among healthcare workers in the gynecology-obstetrics department, May 2024 (*n*=41).

### COVID-19 vaccination

Multivariate analysis showed that healthcare workers with tertiary education were 44 times more likely than those with secondary education not to have been vaccinated against COVID-19, and this was significant (p=0.043). In addition, although not significant, healthcare workers with ICP training were seven times more likely not to be vaccinated against COVID-19 (Table 5).

**Table 5.** Univariate and multivariate analysis of factors associated with non-compliance with COVID-19 vaccination among healthcare workers in the gynecology-obstetrics department, May 2024 (*n*=41)

| Factor | Vaccination<br>*n* (%) |  | Total<br>*n* | cOR | *p*-value | aOR | 95% CI limits |  | *p*-value<sup>2</sup> |
| --- | --- | --- | --- | --- | --- | --- | --- | --- | --- |
|  | No<br>31 (76) | Yes<br>10 (24) |  |  |  |  | Lower | Upper |  |
| **Age<sup>1</sup>** |  |  |  |  |  |  |  |  |  |
| □25 | 12 (80) | 3 (20) | 15 | 1 |  |  |  |  |  |
| 25 - 34 | 7 (78) | 2 (22) | 9 | 0.9 | 0.896 |  |  |  |  |
| 35 - 44 | 2 (40) | 3 (60) | 5 | 0.2 | 0.109 |  |  |  |  |
| 45 + | 10 (83) | 2 (17) | 12 | 1.3 | 0.824 |  |  |  |  |
| **Gender** |  |  |  |  |  |  |  |  |  |
| Female | 24 (75) | 8 (25) | 32 | 1 |  | 1 | 1 |  |  |
| Male | 7 (78) | 2 (22) | 9 | 1.2 | 0.864 | 0.1 | 0.0 | 1.7 | 0.140 |
| **Marital status** |  |  |  |  |  |  |  |  |  |
| Married | 9 (75) | 3 (25) | 29 | 1 |  | 1 | 1 |  |  |
| Single | 22 (76) | 7 (24) | 12 | 1.1 | 0.953 | 0.5 | 0.0 | 5.4 | 0.593 |
| **Educational level** |  |  |  |  |  |  |  |  |  |
| Secondary | 6 (55) | 5 (45) | 11 | 1 |  | 1 | 1 |  |  |
| Higher | 25 (83) | 5 (17) | 30 | 4.2 | 0.066 | 43.6 | 1.4 | 5.2 | **0.043** |
| **Grade** |  |  |  |  |  |  |  |  |  |
| Care assistant | 2 (67) | 1 (33) | 3 | 1 |  | 1 | 1 |  |  |
| Nurse | 9 (69) | 4 (31) | 13 | 1.1 | 0.931 | 0.6 | 0.0 | 27.6 | 0.793 |
| Resident | 6 (75) | 2 (25) | 8 | 1.5 | 0.783 | 0.1 | 0.0 | 6.8 | 0.264 |
| Midwife/Maieutician | 5 (71) | 2 (29) | 7 | 1.3 | 0.880 | 0.1 | 0.0 | 24.0 | 0.346 |
| Medical student | 9 (90) | 1 (10) | 10 | 4.5 | 0.351 | 0.2 | 0.0 | 23.4 | 0.479 |
| **Professional group** |  |  |  |  |  |  |  |  |  |
| Paramedical | 16 (70) | 7 (30) | 13 | 1 |  |  |  |  |  |
| Student | 9 (90) | 1 (10) | 10 | 3.9 | 0.232 |  |  |  |  |
| Medical | 6 (75) | 2 (25) | 8 | 1.3 | 0.770 |  |  |  |  |
| **Refresher course received on ICP measures** |  |  |  |  |  |  |  |  |  |
| Yes | 8 (57) | 6 (43) | 14 | 1 |  | 1 | 1 |  |  |
| No | 23 (85) | 4 (15) | 27 | 4.3 | 0.056 | 7.3 | 0.7 | 122 | 0.122 |
| **Professional experience 1<sup>1</sup>** |  |  |  |  |  |  |  |  |  |
| □5 | 18 (82) | 4 (18) | 22 | 1 |  |  |  |  |  |
| 5 + | 13 (68) | 6 (37) | 19 | 0.5 | 0.324 |  |  |  |  |
| **Professional experience 2<sup>1</sup>** |  |  |  |  |  |  |  |  |  |
| □10 | 19 (79) | 5 (21) | 24 | 1 |  |  |  |  |  |
| 10 + | 12 (71) | 5 (29) | 17 | 0.6 | 0.530 |  |  |  |  |

<sup>1</sup>Unit in years; <sup>2</sup> Fisher exact probability test; cOR: crude Odds Ratio; aOR: adjusted Odds Ratio; CI: Confidence interval

The Johnson & Johnson vaccine was the vaccine most frequently requested by staff who had received the COVID-19 prophylaxis (70%), followed by the Sinopharm vaccine (30%) (Figure 4)

**Fig. 4.**
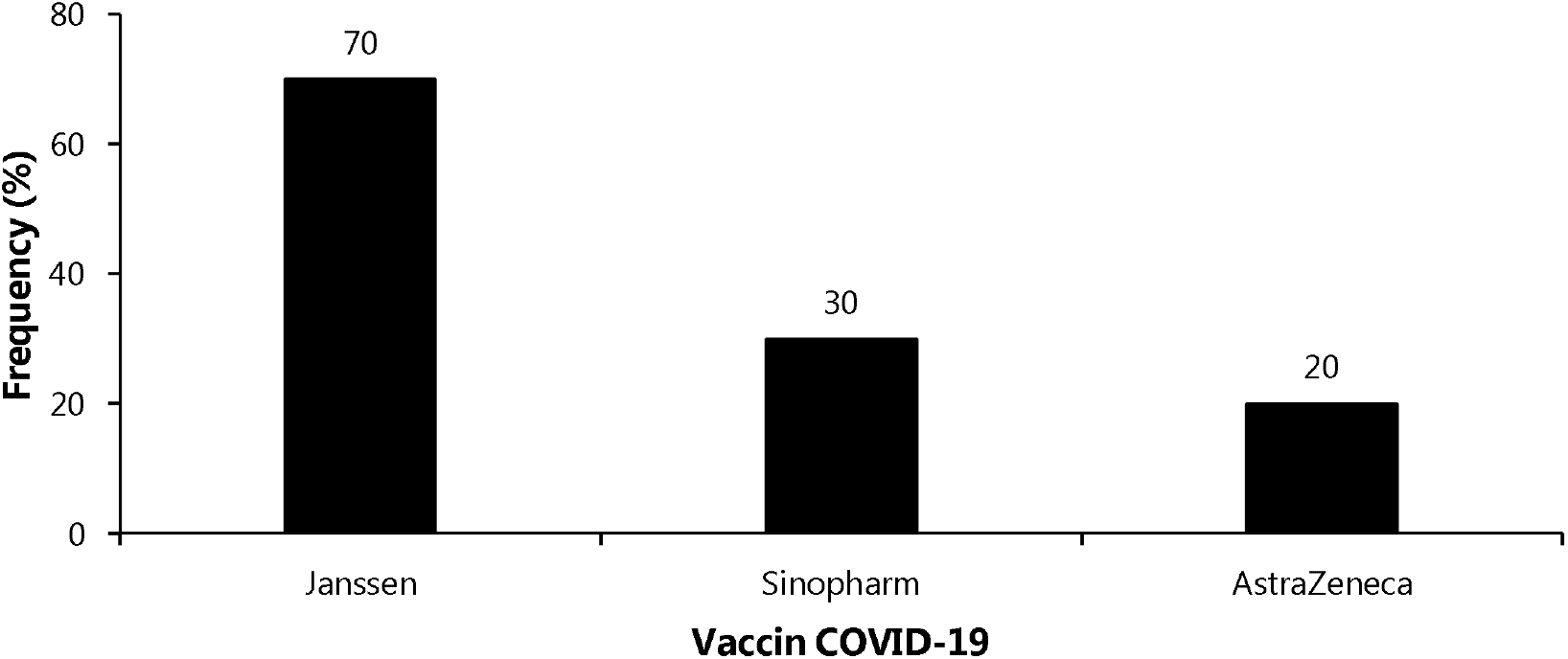
Type of COVID-19 vaccines received by healthcare workers in the gynecology-obstetrics department, May 2024 (*n*=10)

## Discussion

### Knowledge on hand hygiene

The study revealed that less than one-third of HCWs had adequate knowledge of the minimum duration of efficacy of the hydroalcoholic gel. This is due to a lack of awareness of the need for prolonged application of the gel to ensure effective elimination of germs from the hands. This lack of awareness leads to ineffective use of the gel, which favors the persistence of germs and their transmission to patients. In contrast, medical staff (residents) (63%) demonstrated greater knowledge on this topic, reflecting a higher medical culture compared to lower-level health workers. Similar studies in Pakistan also found that medical staff (66%) had better knowledge of this indicator than paramedics [29].

### Perception of hand hygiene

Most of respondents reported that they always practice hand hygiene and consider it an essential part of their care. This is encouraging for HCWs and patients alike, as a correct perception of the importance of hand hygiene is essential in the prevention of HAIs. Despite this good perception of hand hygiene, study reported that HCWs do not always comply with hand hygiene [24,30]. This poor performance has been attributed to the unavailability of wash facilities close to the point of care, the perception that patient care is safe, and high workloads as described in studies conducted in Yaounde health districts and the Democratic Republic of Congo [24,31].

The study reports emphasize that suboptimal adherence to universal precautions indicate inadequate supervision of healthcare facilities and their staff by intermediate or strategic levels [30]. In such circumstances, the role of infection control committees and internal and external monitoring guidelines is critical in promoting compliance. Measures to improve accessibility and availability of laundry facilities, reduce staff workload, and increase supervision and training are essential to ensure optimal adherence to universal precautions and reduce the risk of healthcare associated infections [30,32].

Hand hygiene is recognized as the single most important measure for preventing the spread of (HAIs) in healthcare facilities. Current standards state that to promote hand hygiene compliance, the necessary resources must be readily available in critical locations and that behavior change must be supported by education, training, monitoring, feedback and organizational support. Research supporting this approach also indicates that to effectively interrupt the transmission of healthcare-associated pathogens, hand hygiene must be performed at the times and places where transmission is most likely to occur. Educating healthcare workers about effective hand hygiene techniques and compliance is therefore essential. Assessing hand hygiene knowledge, perceptions and practices helps to build capacity in this area [2,33].

### Accidental exposure to blood and other body fluids

More than half of the respondents (59%) experienced an accidental exposure to biological fluids in the past 12 months. Splashes were the main source of exposure (96% of cases). Our results confirmed the trends observed in previous studies conducted at the Yaounde university teaching hospital and in the Yaounde Health Districts [9,10]. However, they were lower than the proportion of exposure recorded at the referral hospitals in Buea and Bertoua, which reported prevalence rates of over 80% [8,34]. Factors related to the healthcare environment may explain these differences.

Blood and amniotic fluid were the biological fluids most frequently involved in occupational exposures of HCWs. In maternity wards, childbirth is a major source of blood and amniotic fluid splashes, as reported in other studies in Cameroon, Côte d’Ivoire, and Morocco [24,35,36].

Exposures from splashes are common and affect most of HCWs [24,34,36]. To manage them, it is essential to use PPE systematically and correctly, and to be immunized against diseases transmitted by biological fluids, such as viral hepatitis B and COVID-19 [9,24]. Although HCWs are aware of these infections, the uptake of preventive vaccination against viral diseases remains insufficient in Cameroon and the rest of the world [9,12,14,24,37,38].

Multivariate analysis of the parameters associated with accidental exposure to biological fluids showed that healthcare workers with a higher level of education were 14 times more likely to be exposed than those with a secondary level of education. Workers with a higher level of education were more likely to be involved in emergency care or surgical procedures, which are the main sources of occupational exposure to blood. Similar findings were made in Cameroon, Ethiopia and Iran [39–42].

### Healthcare workers adherence to preventive vaccination

The coverage of fully vaccinated HCWs was 27% for viral hepatitis B and 19% for COVID-19. However, no participants were fully vaccinated against cholera. HCWs cite several reasons for the low uptake of vaccination, including the high cost of the vaccine, its unavailability, lack of time, doubts about its content and fear of side effects, etc. [9,24,38]. This low vaccination coverage makes healthcare workers particularly vulnerable to life-threatening infectious diseases such as viral hepatitis, cholera and COVID-19, and facilitates their transmission to other and patients and HCWs.

Half of HCWs exposed to blood and body fluids were not fully vaccinated against viral hepatitis B. This alarming vulnerability increases the likelihood of seroconversion among HCWs and highlights a critical missed opportunity for preventive vaccination and consequent reduction in healthcare costs. To address this issue, policymakers and healthcare administrators should consider introducing mandatory pre-employment hepatitis B vaccination for HCWs or implementing employer-funded hepatitis B vaccination programs for HCWs [9]. By adopting these strategies, health institutions can substantially mitigate occupational transmission of hepatitis B, protect the health of HCWs, and reduce the financial burden on the healthcare system.

The Johnson & Johnson vaccine was the most commonly requested vaccine by workers who had received the preventive vaccine against COVID-19. Our observations confirmed the trends observed in the Cameroonian population in general, and specifically among HCWs [24,43].

Analysis of the determinants of vaccination showed that HCWs with higher levels of education or who had not received training in ICP measures were less likely to be vaccinated. These findings raise the question of whether HCWs with higher levels of education may be more confident in their ability to avoid infection and less likely to be vaccinated. Similarly, a lack of training in ICP interventions could lead to a lack of awareness of the risks and benefits of vaccination. These findings suggest that it is essential to set up awareness and training programs to improve knowledge of the infectious risks and benefits of vaccination and to encourage HCWs to be vaccinated.

## Limitations

The main limitation of this study is the risk of recall bias regarding previous exposure, or the difficulty in proving that exposure to the risk was prior to the outcome. In addition, the sample size did not provide sufficient power to detect statistical significance for some variables that were not considered significant. These limitations highlight the need for a more comprehensive study with a larger sample size.

## Conclusions

This study shows that inadequate knowledge of hand hygiene, high occupational exposure to biological fluids and low vaccination coverage make HCWs in this ward particularly vulnerable to infectious diseases. This also increases the risk HAIs to women and newborns. Healthcare facility managers and national health authorities must therefore commit to implementing specific strategies targeted at maternity ward staff. These strategies should include increasing staff training in standard precautions and promoting vaccination among HCWs.

## Data Availability

All data produced in the present work are contained in the manuscript

## Abbreviations

aOR: Adjusted Odds Ration
CI: Confidence Interval
cOR: Crude Odds Ratio
COVID-19: New Coronavirus Disease
HAI: Healthcare Associated Infection
HCW: Healthcare Worker
ICP: Infection Control and Prevention

## Declaration

### Authors’ contribution

Drafting of the study protocol, data collection, analysis, interpretation, drafting and editing of manuscript: F.Z.L.C.

### Ethical approval statement

The protocol was approved by Institutional Review Board (IRB) of the Faculty of Medicine and Biomedical Sciences of Yaounde and the ethical clearance: N°1017/UYI/FMSB/VDRC/DAASR/CSD issued. Informed consent was obtained from participants prior to inclusion in the study. All methods were performed according to relevant guidelines and regulations.

### Consent for publication

Not applicable.

### Availability of data and materials

All data generated or analyzed during this study are included in this published article.

## Acknowledgements

Our gratitude goes to the health staff who agreed to participate in this study and to the manager of the health facility who gave an authorization to conduct the study.

